# One year cross-sectional study in adult and neonatal intensive care units reveals the bacterial and antimicrobial resistance genes profiles in patients and hospital surfaces

**DOI:** 10.1101/2020.02.11.20022145

**Authors:** Ana Paula Christoff, Aline Fernanda Rodrigues Sereia, Giuliano Netto Flores Cruz, Daniela Carolina de Bastiani, Vanessa Leitner Silva, Camila Hernandes, Ana Paula Metran Nascente, Ana Andrea dos Reis, Renata Gonçalves Viessi, Andrea dos Santos Pereira Marques, Bianca Silva Braga, Telma Priscila Lovizio Raduan, Marines Dalla Valle Martino, Fernando Gatti de Menezes, Luiz Felipe Valter de Oliveira

**Affiliations:** BiomeHub, Florianopolis, Brazil; Albert Einstein Israelite Hospital, São Paulo, Brazil

**Keywords:** Hospital microbiome, 16S rRNA, bacteria, antimicrobial resistance, high throughput sequencing, MDR, HAI

## Abstract

Several studies have shown the ubiquitous presence of bacteria in the hospital environmental surfaces, staff, and patients. Frequently, these bacteria are related to HAI (healthcare-associated infections) and carry antimicrobial resistance (AMR). These HAI-related bacteria contributes to a major public health issue by increasing patient morbidity and mortality during or after hospital stay. Bacterial high-throughput amplicon gene sequencing along with AMR genes identification and whole genome sequencing (WGS) are new biotechnological tools that allow multiple-sample screening for a diversity of bacteria. In this paper, we used these methods to perform an one-year cross sectional profiling of bacteria and AMR genes in adult and neonatal intensive care units (ICU and NICU) in a Brazilian public hospital. Our results showed high abundances of HAI-related bacteria as *S. epidermidis, S. aureus, K. pneumoniae, A. baumannii* complex, *E. coli, E. faecalis* and *P. aeruginosa* in patients and hospital surfaces. Most abundant AMR genes detected throughout ICU and NICU were *mecA, bla*_CTX-M-1 group_, *bla*_SHV-like_ and *bla*_KPC-like_. We found that NICU environment and patients were more widely contaminated with pathogenic bacteria than ICU. Patient samples, despite the higher bacterial load, have lower bacterial diversity than environmental samples in both units. Finally, we also identified contamination hotspots in the hospital environment showing constant frequencies of bacterial and AMR contamination throughout the year. WGS sequencing, 16S rRNA oligotypes and AMR identification allowed a high-resolution data analysis for bacterial characterization and its distributions along the hospital microbiome profile.

## Introduction

Hospital indoor environment including surfaces, staff and patients are the focus of major recent investigations regarding the microbiome and its bacterial composition (1–4). This became possible due to the dissemination of culture-independent microbiology methods using next-generation DNA sequencing. Culture-independent methods are less time-consuming and skewed to specific cultivable microorganisms, allowing large-scale screening and surveillance of microorganisms, including those that do not grow well in laboratory conditions (5–9).

Several studies already demonstrated the presence and persistence of bacterial pathogens in hospital surfaces, like *Enterococcus, Staphylococcus aureus, Acinetobacter, Escherichia coli, Klebsiella, Pseudomonas aeruginosa* and *Serratia marcescens* (10–12). These microorganisms, among others listed by CDC – Centers for Disease Control and Prevention (https://www.cdc.gov/hai/index.html) - are known as nosocomial pathogens related to Healthcare-associated infections (HAI). These pathogenic bacteria can also harbor multidrug-resistance genes (MDR) and confer a broad spectrum of antimicrobial resistance (AMR) - a matter of major concern in public health, leading to increased patient morbidity and mortality during or after hospital stay (13,14).

Hospital contaminated environments, including surfaces and staff, are well recognized as reservoirs and transmission sources for HAI-related pathogens (2,12,15–21). In this scenario, the study of hospital microbiome has substantial implications in the healthcare system: the continuous monitoring of hospital environment, bacterial tracking, and detailed epidemiological investigations may contribute to decrease HAI rates and improve healthcare system quality. In this study, by monthly collecting DNA samples from environment and patients, we investigated the microbiome of adult and neonatal intensive care units (ICU and NICU) from a local public hospital over a one-year period. We perform exhaustive molecular assessment to understand the profiles of bacterial abundances as well as antimicrobial resistance genes from the healthcare environment and patient samples.

## Material and Methods

### Study design

This study was performed in a public hospital in the city of São Paulo, Brazil. A twelve-month project was designed comprising adult intensive care unit (ICU) and neonatal intensive care unit (NICU) samples from environmental surfaces and patients. From August-2018 to July-2019, patient beds and common use areas were sampled each month, as well as patient nasal and rectal swabs. For neonatal patients, stool swabs were collected instead of rectal. Bed environmental surface samples were collected from medical and hospital equipment, furniture, critical structure points, and bed accessories – all characterized as high contact surfaces (Supplementary table 1). All patient samples collected were from the same beds included in the environmental sampling. Among common areas included are: nurse station, as well as common use medical equipment. Additionally, some bacterial isolates were obtained by the hospital microbiology laboratory for further genomic analysis. This project was approved by the Albert Einstein Israelite Hospital ethics committee (number 2.585.209) All participants were informed about the study aims and sampling was carried out upon a signature of an informed consent by the patient or a legal representative.

**Table 1.** Microorganisms isolated in the hospital microbiology laboratory in parallel to the project development (Aug-2018 to July-2019) and the subsequent additional identifications using 16S rRNA V3/V4 amplicon sequencing and whole genome sequencing (WGS) by the molecular biology laboratory.

| Month | Unit | Microbiology Lab identification | 16S rRNA -V3/V4 identity | 16S rRNA oligotype | WGS identity | MLST identity | Clonal group identity |
| --- | --- | --- | --- | --- | --- | --- | --- |
| N/A* | N/A | *Staphylococcus aureus* | *Staphylococcus aureus* | oligotype_4 | *Staphylococcus aureus* | ST1435 | - |
| Oct-18 | NICU | *Staphylococcus aureus* | *Staphylococcus aureus* | oligotype_4 | *Staphylococcus aureus* | ST2383 | *S. aureus* Group 1 |
| Oct-18 | NICU | *Staphylococcus aureus* | *Staphylococcus aureus* | oligotype_4 | *Staphylococcus aureus* | ST398 | *S. aureus* Group 2 |
| Oct-18 | NICU | *Staphylococcus aureus* | *Staphylococcus aureus* | oligotype_4 | *Staphylococcus aureus* | ST2383 | *S. aureus* Group 1 |
| Jan-19 | NICU | *Staphylococcus aureus* | *Staphylococcus aureus* | oligotype_4 | *Staphylococcus aureus* | ST398 | *S. aureus* Group 2 |
| Nov-18 | ICU | *Staphylococcus epidermidis* | *Staphylococcus epidermidis* | oligotype_1 | *Staphylococcus epidermidis* | ST2 | - |
| Sep-18 | ICU | *Enterococcus faecalis* | *Enterococcus faecalis* | oligotype_5 | *Enterococcus faecalis* | ST21 | - |
| Feb-19 | NICU | *Enterococcus faecalis* | *Enterococcus faecalis* | oligotype_5 | *Enterococcus faecalis* | ST21 | - |
| Dec-18 | ICU | *Pseudomonas aeruginosa* | *Pseudomonas aeruginosa* | oligotype_12 | *Pseudomonas aeruginosa* | ST245 | - |
| Feb-19 | NICU | *Pseudomonas aeruginosa* | *Pseudomonas aeruginosa* | oligotype_12 | *Pseudomonas aeruginosa* | ST1816<sub>1</sub> | - |
| Oct-18 | ICU | *Klebsiella* | *Klebsiella pneumoniae* | - | *Klebsiella pneumoniae* | ST307 | - |
| Oct-18 | NICU | *Klebsiella oxytoca* | *Enterobacteriaceae* | - | *Klebsiella michiganensis* | N/A | - |
| Nov-18 | ICU | *Escherichia coli* | *Escherichia coli* | oligotype_2 | *Escherichia coli* | ST131 | - |
| Jan-19 | NICU | *Escherichia coli* | *Escherichia coli* | - | *Escherichia coli* | ST69 | *E. coli* Group 1 |
| Jan-19 | NICU | *Escherichia coli* | *Escherichia coli* | - | *Escherichia coli* | ST69 | *E. coli* Group 1 |
| Oct-18 | ICU | *Escherichia coli* | *Escherichia coli* | oligotype_2 | *Escherichia coli* | ST410 | - |
| Sep-18 | ICU | *Escherichia coli* | *Escherichia coli* | oligotype_2 | *Escherichia coli* | ST10 | - |

\*Isolated in the hospital previously the project period.
<sub>1</sub> Has one allele with less than 100% identity (99.7992%).

### Sample collection and DNA extraction

Samples were collected using a sterile dry cotton swab, without transport media. The swab was moistened with a sterile saline solution (0.9% NaCl) prior to sample collection. The swabs were transported to the laboratory facilities at room temperature and processed within a maximum of 24h after sample collection. Bacterial DNA from the samples was obtained through a thermal lysis (96 °C – 10 min) followed by a purification step with AMPure XP Magnetic Beads (Beckman Coulter, USA). Negative controls were included in each lysis and DNA extraction batch.

### Library preparation and DNA sequencing

We performed amplicon sequencing library preparation for bacteria using the V3/V4 16S rRNA gene primers 341F and 806R (22,23) in a two-step equivolumetric PCR protocol (24). The first PCR was performed with V3/V4 universal primers containing a partial Illumina adaptor, based on TruSeq structure (Illumina, USA) that allows a second PCR with the indexing sequences similar to procedures described previously (25). Here, we add unique dual-indexes per sample in the second PCR. Two microliters of individual sample DNA were used as input in each first PCR reaction. The PCR reactions were carried out using Platinum Taq (Invitrogen, USA) with the conditions: 95°C for 5 min, 25 cycles of 95°C for 45s, 55°C for 30s and 72°C for 45s and a final extension of 72°C for 2 min for PCR 1. In PCR 2 the conditions were 95°C for 5 min, 10 cycles of 95°C for 45s, 66°C for 30s and 72°C for 45s and a final extension of 72°C for 2 min. All PCR reactions were performed in triplicate. The final PCR reactions were cleaned up using AMPureXP beads (Beckman Coulter, USA) and an equivalent volume of each sample was added in the sequencing pool. At each batch of PCR, a negative reaction control was included (CNR). The final DNA concentration of the libraries pool was estimated with Picogreen dsDNA assays (Invitrogen, USA) and then diluted for accurate qPCR quantification using KAPA Library Quantification Kit for Illumina platforms (KAPA Biosystems, USA). The sequencing pool was adjusted to a final concentration of 11 pM (for V2 kits) or 18 pM (for V3 kits) and sequenced in a MiSeq system (Illumina, USA), using the standard Illumina primers provided by the manufacturer kit. Single-end 300 cycle runs were performed using V2×300, V2×500 or V3×600 sequencing kits (Illumina, USA) with sample coverages set to 45,000 reads per sample in all sequencing runs (Supplementary table 2).

### Sequencing data analysis

The sequenced reads obtained were processed using an *in-house* developed bioinformatics pipeline described below (BiomeHub, Brazil – hospital_miccrobiome_rrna16s:v0). Illumina FASTQ files had the primers trimmed and their accumulated error evaluated (24). Only one mismatch is allowed in the primer sequence that should be present at the beginning of the read. The whole read sequence is discarded if this criterion is not met. Reads were analyzed with the Deblur package (26) to remove possible erroneous reads and then identical read sequences were grouped into oligotypes (clusters with 100% identity). The sequence clustering with 100% identity provides a higher resolution for the amplicon sequencing variants (ASVs), also called sub-OTUs (sOTUs) (27) - herein denoted as oligotypes. Next, VSEARCH (28) was used to remove chimeric amplicons.

An additional filter was implemented to remove oligotypes below the frequency cutoff of 0.2% in the final sample counts. We also implemented a negative control filter, since hospital microbiome generally are low biomass samples (24). In each processing batch, we also used negative controls for the DNA extraction and PCR reactions. If any oligotypes are recovered in the negative control results, they are checked against the samples and automatically removed from the results only if their abundance (in number of reads) are no greater than two times their respective counts in the sample. The remaining oligotypes in the samples are used for taxonomic assignment with the BLAST tool (29) against a reference genomic database (encoderef16s_rev6_190325). This *in-house* database was constructed with complete and draft bacterial genomes, focused on clinically relevant bacteria, obtained from NCBI. It is composed of 11,750 sequences including 1,843 different bacterial taxonomies.

Taxonomy was assigned to each oligotype using a lowest common ancestor (LCA) algorithm. If more than one reference can be assigned to the same oligotype with equivalent similarity and coverage metrics (e.g. two distinct reference species mapped to oligotype “A” with 100% identity and 100% coverage), the taxonomical assignment algorithm leads the taxonomy to the lowest level of possible unambiguous resolution (genus, family, order, class, phylum or kingdom), according to the similarity thresholds previously established (30).

After quality check of sequencing yield (Supplementary Table 2), the resulting oligotype tables, analogous to traditional OTU tables, were processed and normalized as previously described (24). Oligotype sequences served as input for FastTree 2.1 software (31) to construct phylogenetic trees and allow beta-diversity analysis with UniFrac distances (32). Additional analysis were performed using R (version 3.6.0) and the Phyloseq package (33). When suited, non-parametric comparisons were performed using Kruskall-Wallis and Wilcoxon tests implemented in R (34). Alpha diversity analysis was performed using the “plot_richness” function in Phyloseq.

### RGene – antimicrobial resistance gene analysis

A panel comprising relevant β-lactamases, Vancomycin, Methicillin and Colistin antimicrobial resistance genes in Brazilian scenario was tested: *bla*_CTX-M-1_ group, *bla*_CTX-M-2_ group, *bla*_CTX-M-8_ group, *bla*_CTX-M-9_ group, *bla*_GES-like_, *bla*_IMP-like_, *bla*_KPC-like_, *bla*_NDM-like_, *bla*_SHV-like_, *bla*_SPM-like_, *bla*_VIM-like_, *bla*_OXA-143-like_, *bla*_OXA-23-like_, *bla*_OXA-48-like_, *bla*_OXA-51-like_, *bla*_OXA-72-like_, *vanA, vanB, mecA* and *MCR1*. The detection was performed using Real-Time PCR with QSY hydrolysis probes labeled with FAM®, VIC® and NED® (Applied Biosystems, USA).

Real Time PCR reactions were carried out using 10 μL of final volume per sample, containing 2 μL of the same previously sequenced DNA samples, 0.2 U Platinum Taq, 1 X Buffer, 3 mM MgCl2, 0.1 mM dNTP, 0.12 X ROX™ and 0.2 μM of each forward and reverse specific primer following the thermal conditions: 95 °C for 5 min with 35 cycles of 95 °C for 15s, 60 °C for 30s and 72°C for 30s. Negative and positive reaction controls were included in all the assays. All the samples were analyzed in experimental triplicate. Real Time reactions were performed in an ABI 7500 Fast Real-Time PCR System (Applied Biosystems, USA). Samples were considered positive when at least two of the experimental replicates were below the quantification cycle 33 using an experimental threshold of 0.05.

### Bacterial genomes

Bacterial isolates were processed as described above for DNA extraction, and the sequencing library preparation was performed using Nextera Flex with CD indexes (Illumina, USA), according to manufacturer instructions. Samples were sequenced in a MiSeq system (Illumina, USA), in paired-end 150 pb configuration, with coverage of 1 million reads per sample.

Sequenced genomes were analyzed using A5 (35), SPAdes (36) and Prokka (37). Species identification and average nucleotide identity (ANI) were performed using the JSpecies platform (38). The whole genome sequence was used to identify the Multi Locus

Sequence Typing (MLST 2.0) as previously described (39). Clonality analysis was performed among bacteria of same species using NDTree (40) to assess genome single nucleotide variants (SNVs) and evaluating ANI values using JSpecies ANIm calculations (38).

## Results

### Samples and high-throughput amplicon sequencing

A total of 1,978 samples were collected from August-2018 to July-2019. In the adult intensive care unit (ICU) 1,062 samples were collected, of which 138 were from patients, 676 from beds, and 248 from the common healthcare environment. In the neonatal intensive care unit (NICU), 916 samples were collected, being 111 from patients, 598 from beds, and 207 from common healthcare environment (detailed sample collection sites described in Supplementary Table 1).

Twelve sequencing runs were performed resulting in 78,840,894 reads, with an average and standard deviation of 6,570,075 ± 813,906 reads per run and 30,942 ± 3,468 reads by sample (Supplementary table 2). In the log^10^ scale, patient and healthcare environment median reads in ICU and NICU reflects the scales of microbial load in the samples (Figure 1). Some months showed lower medians, suggesting samples with lower bacterial load, however, the annual profile is pretty similar.

**Figure 1.**
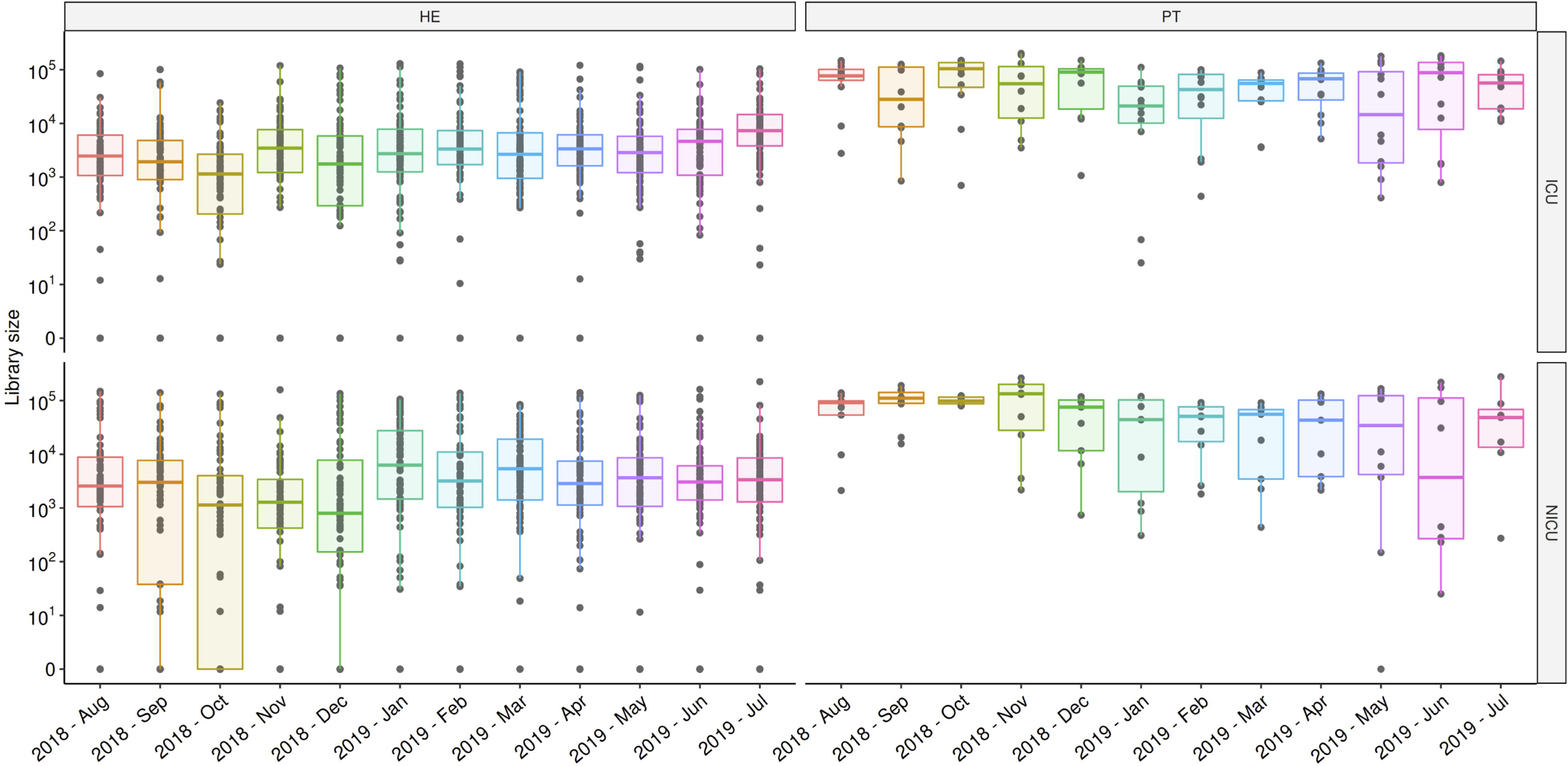
Total sequenced reads (library size) for each month of samples collected in ICU and NICU healthcare environment (HE) and patients (PT). Boxplots represents median distributions for the total reads obtained in samples, considering log^10^ scales.

Considering quality reads that passed through our bioinformatics pipeline, 98.74% of reads could be classified as bacteria (kingdom), 97.84% were classified at family level, 89.14% at genus level, and 67.28% at species level. Of the global sequencing results, 40.96% of the reads could be identified in a HAI-related Bacteria group (here denoted HAIrB group: *Acinetobacter baumanii* complex, *Escherichia coli, Enterococcus faecalis, Enterococcus faecium, Klebsiella pneumoniae, Proteus mirabilis, Pseudomonas aeruginosa, Staphylococcus aureus* and *Staphylococcus epidermidis)*, corresponding to 60.87% of the results at species level.

Throughout the assessed year, 19.79% of the identified bacterial DNA belonged to the genus *Staphylococcus* (11.79% *S. epidermidis* and 3.03% *S. aureus*, 4.97% other species). Other HAIrB had the following frequencies: 7.39% - *A. baumannii* complex, 7.06% - *E. coli*, 3.27% - *Enterococcus* and 5.73% - *K. pneumoniae*. Thus, we can state that the genus *Staphylococcus* and the species *S. epidermidis* have predominant observed proportions in this hospital microbiome throughout this year of analysis.

### Bacterial diversity analysis

Alpha diversity analysis for both ICU and NICU showed higher and more variable values of Shannon index in the healthcare environment than in patient samples along the year (Figure 2a). Similar pattern was observed when looking at specific sampling sites within units (Figure 2b). Longitudinal profiles of alpha diversity along the year in different hospital locations and specific collection sites are shown in Supplementary figure 1, revealing highly variable values even among equivalent sample sites within and between months.

**Figure 2.**
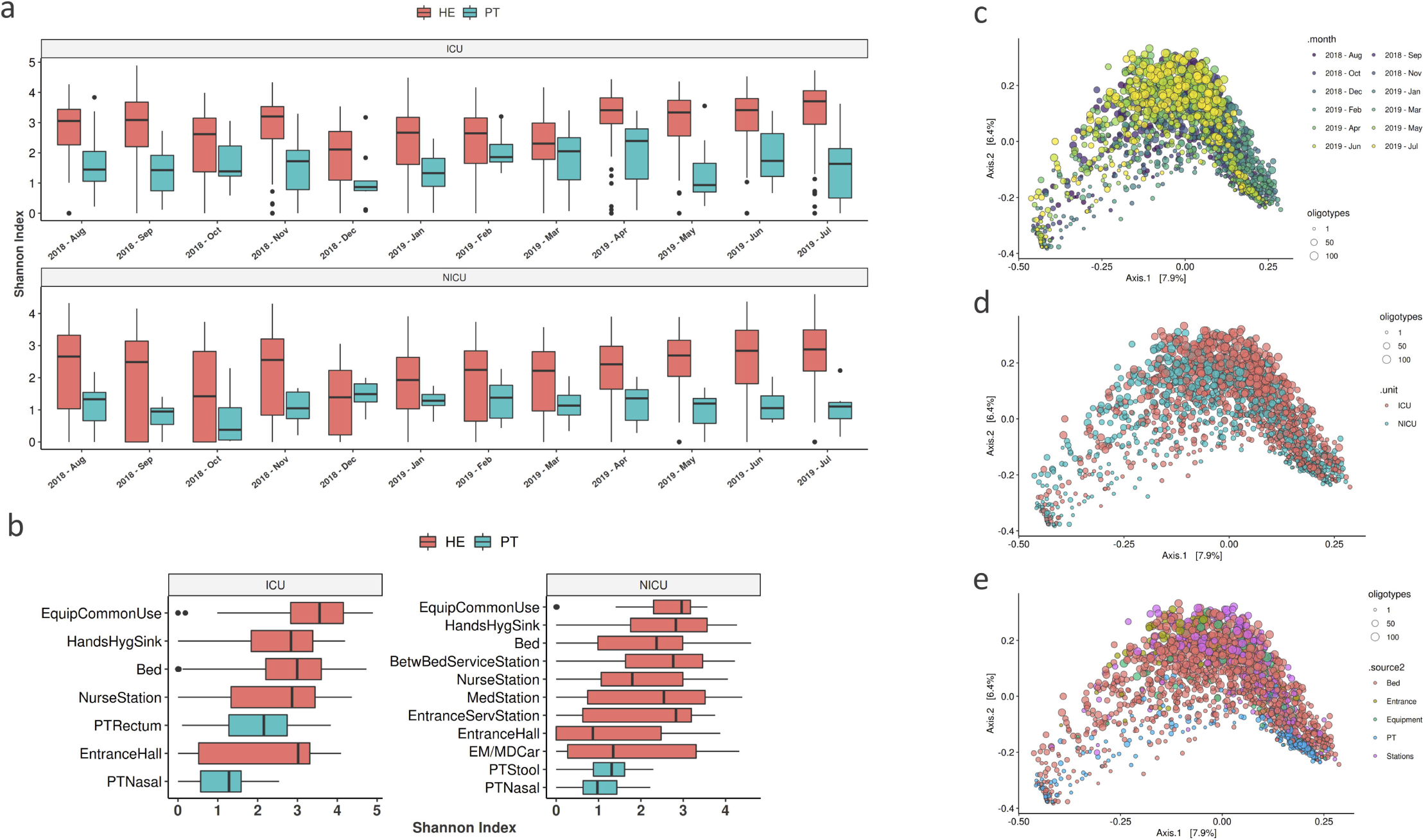
Diversity analysis. **(A)** Shannon alpha-diversity metrics for ICU and NICU healthcare environment (HE) or patient (PT) samples along the year of study, **(B)** Shannon index calculated for sample collected locations. PCoA plots with beta-diversity weighted UniFrac analysis considering the samples groups by **(C)** Month - August-2018 to Jul-2019, **(D)** Unit - ICU and NICU and **(E)** Collection site: Bed, Entrance, Equipment, Patient (PT) and Stations.

Weighted UniFrac analysis showed similar distribution patterns among samples across the year (Figure 2c), without clear separation between months, but with a slightly differential distribution of samples from August to November 2018, December 2018 to February 2019, and April to July 2019 (Supplementary figure 2). ICU and NICU samples have similar beta-diversity profiles (Figure 2d), and only patient samples seem to cluster more closely (Figure 2e).

### Bacterial profiling in the hospital

Several bacterial taxonomies were identified in this study and classified into phylum, family, genus and species levels. Proteobacteria are by far the most predominant bacteria phylum found in the results of both ICU and NICU, followed by Firmicutes, Actinobacteria and Bacteroidetes. The most abundant families found were: Enterobacteriaceae, Staphylococcaceae, Moraxellaceae, Pseudomonadaceae, Corynebacteriaceae, Streptococcaceae, Enterococcaceae, Peptoniphilaceae, Prevotellaceae and Bacteroidaceae; major genus were: *Staphylococcus, Acinetobacter, Pseudomonas, Corynebacterium, Klebsiella, Escherichia, Streptococcus, Enterococcus, Bacteroides* and *Proteus*. This higher rank classifications are also in agreement with the most representative species found in the data: *Staphylococcus epidermidis, Klebsiella pneumoniae, Acinetobacter baumannii* complex, *Escherichia coli, Staphylococcus aureus, Enterococcus faecalis, Pseudomonas aeruginosa, Acinetobacter iwoffi, Pseudomonas stutzeri* and *Staphylococcus haemolyticus*. The top 10 bacteria found for each taxonomy level in the results from ICU and NICU can be visualized in Supplementary figure 3, faceted by time points and sampling sites.

**Figure 3.**
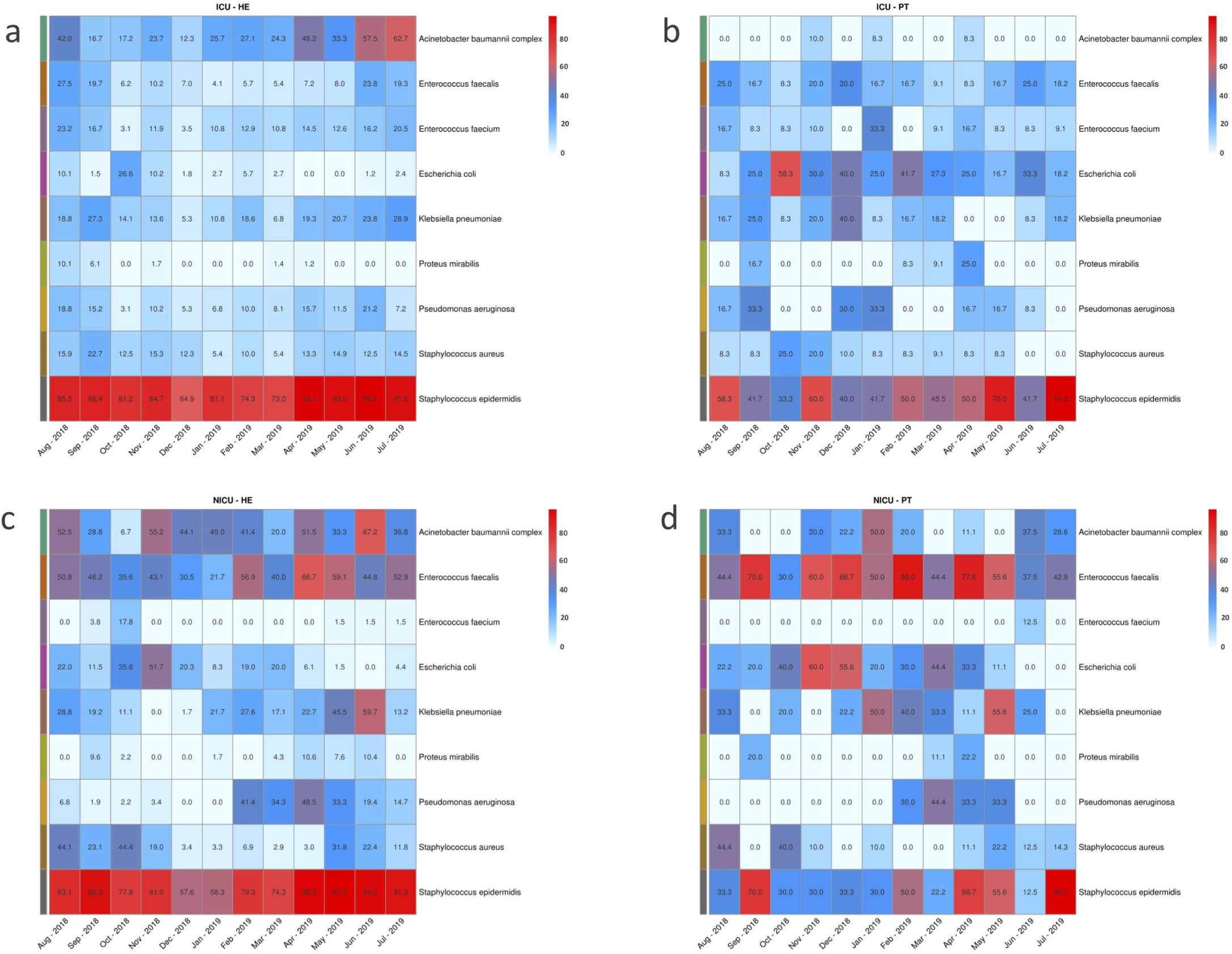
Most abundant bacterial proportion in samples from each month for **(A)** ICU environment, **(B)** ICU patients, **(C)** NICU environment and **(D)** NICU patients. Color scales indicates the most abundant proportions (red) to the least abundant (white).

Considering the previously defined HAIrB group we evaluated the proportion of positive samples for each of the individual HAIrB species in each month of the project. ICU healthcare environment and patients (Figure 3a and 3b) share high proportions of positive samples for *S. epidermidis*. In the environment, *S. epidermidis* was constant all over the year, but patient samples has a more oscillating pattern. *A. baumannii* complex is more present in environment samples, while *E. coli* is proportionally more detected in patient samples. In the overall profile it seems that ICU healthcare environment have constant positivity proportions for the species along the year, where ICU patients varies for most bacteria, not being at constant rates along the year. NICU bacteria detected in most environmental samples are *S. epidermidis, E. faecalis* and *A. baumanii* complex, with higher proportions in samples along the year. Environmental *E. coli, K. pneumoniae*, and *S. aureus* were more temporally distributed, resembling the NICU patient positive samples proportions (Figure 3c and 3d). In patient samples, the bacteria with highest detection rates were *E. faecalis, E. coli, K. pneumoniae*, and *S. epidermidis*.

### Contamination hotspots and longitudinal profiles

Given the observed HAIrB annual profiles, we aimed to identify possible hospital locations as contamination hotspots. Figures 4a through 4f suggest that patient samples show higher total abundance of HAIrB than environmental samples, regardless of the hospital unit. The pattern is consistent even when we split the samples according to sampling sites grouped by similarity (Figures 4b and 4e).

**Figure 4.**
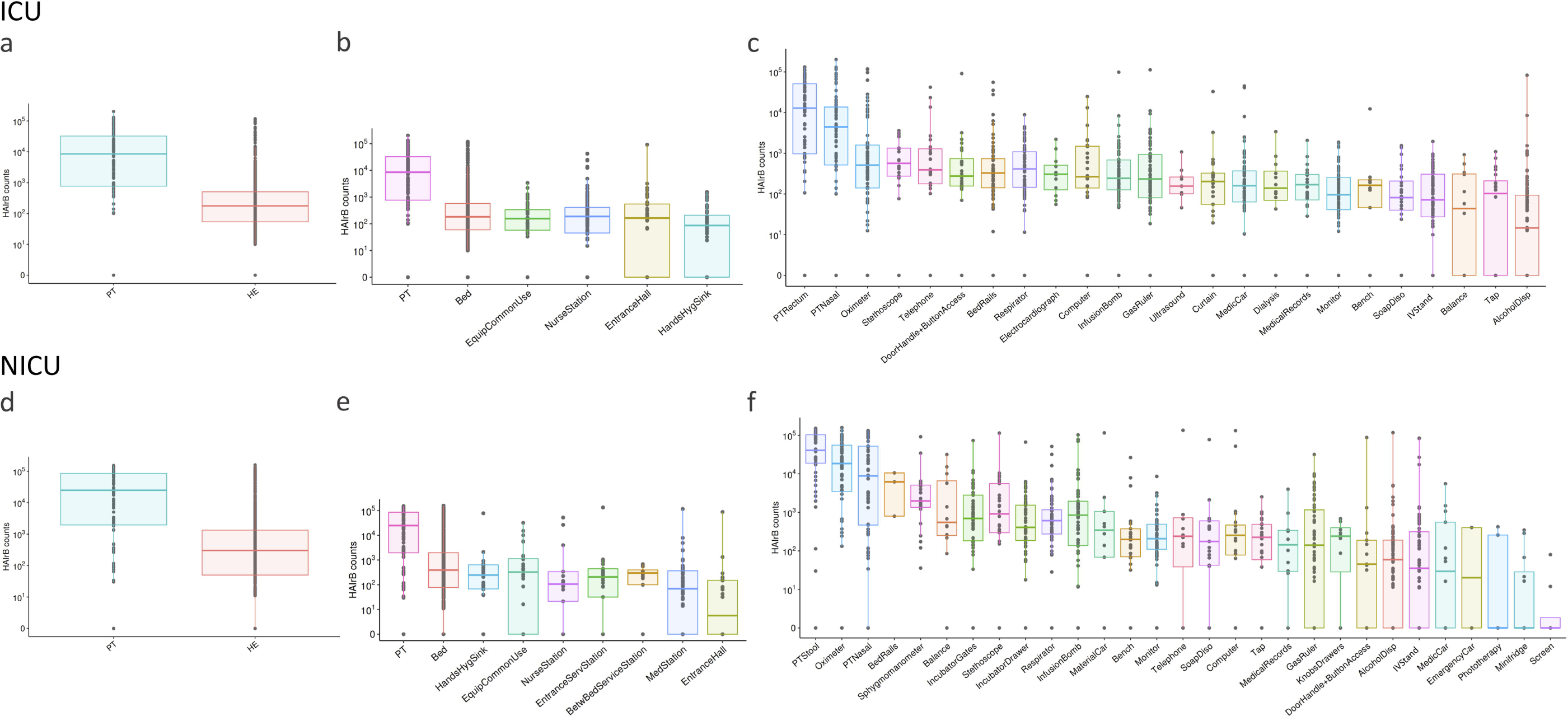
Bacterial sequences detected for HAI related bacteria (HAIrB counts), including *Acinetobacter baumanii* complex, *Escherichia coli, Enterococcus faecalis, Enterococcus faecium, Klebsiella pneumoniae, Proteus mirabilis, Pseudomonas aeruginosa, Staphylococcus aureus* and *Staphylococcus epidermidis* in ICU samples for patient (PT) and healthcare environment (HE) **(A)**, sample locations **(B)** and specific sampling sites **(C)**. HAIrB counts for NICU samples considering patient (PT) and healthcare environment (HE) **(D)**, sample locations **(E)** and specific sampling sites **(F)**.

Within ICU, the highest HAIrB levels were observed in PT samples, with progressively lower values for bed, common use equipment, nurse station, entrance hall, and hands hygiene sink. More specifically, ICU patient rectum and nasal samples have the higher HAIrB values (Figure 4c), while the balances, taps, and alcohol dispensers showed the lowest contamination levels. NICU locations also showed highest HAIrB abundance for PT samples and lowest for entrance hall (Figure 4e). In specific sample collection sites, NICU patient stool, oximeter, and patient nasal samples have the higher HAIrB values (Figure 4f), while door handles plus access buttons, alcohol dispensers, IV stand, medication car, emergency car, phototherapy, minifridge and screen had the lowest HAIrB median counts.

The HAIrB profiles can be assessed with respect to their longitudinal variation (Figures 5a and 5b). While not all sites were evenly sampled, many show consistent trends. For instance, medical equipment such as oximeter, monitor, and infusion bombs showed almost constant abundance over the entire year, regardless of the hospital unit. Some sampling sites such as ICU alcohol dispenser and tap showed higher values at the end of the study period. A similar pattern was observed in NICU gas ruler, although with seemly high initial levels.

**Figure 5.**
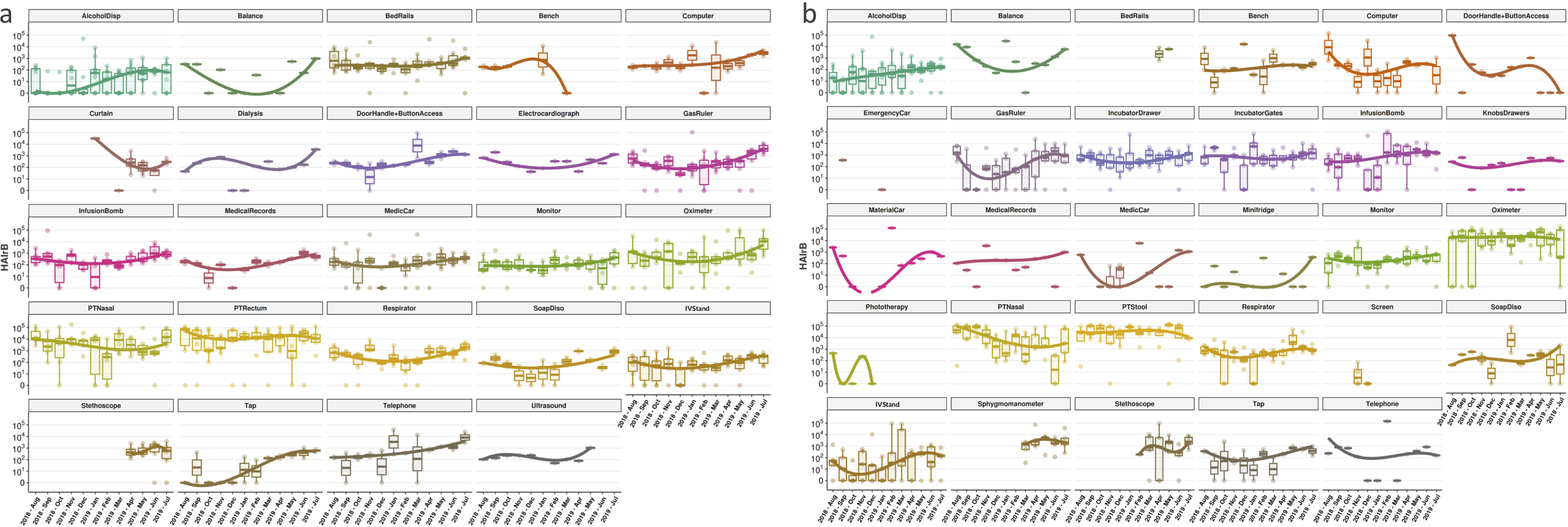
Longitudinal profile of bacteria present in samples specific sites collected along the year for ICU (**A**) and NICU (**B**), considering the previously established HAIrB group: *Acinetobacter baumanii* complex, *Escherichia coli, Enterococcus faecalis, Enterococcus faecium, Klebsiella pneumoniae, Proteus mirabilis, Pseudomonas aeruginosa, Staphylococcus aureus* e *Staphylococcus epidermidis*. Sequence reads are shown in log^10^ scales.

Nonetheless, many sites show heterogeneous profiles in terms of HAIrB contamination. For instance, NICU computer and infusion bombs, as well as ICU telephone and door handle plus access button, show well defined sample distributions outside the trend line in various months. This could be result of occasional increase or decrease in decontamination practices by hospital staff, but also a sampling artifact (especially in those cases in which sample size is minimal, e.g., NICU medical records). Additionally, some sites were not evaluated at all time points. The ICU bench reached particularly low values in March-2019 and was not further monitored, while curtains and stethoscopes started being assessed near the middle of the study period. This is in agreement with the ethical guidelines imposed on this study as monitoring was primarily driven by patient-care concerns.

### Source tracking and bacterial dispersion

Oligotype sequences, corresponding to amplicon sequence variants (ASVs), were used to investigate the bacterial dispersion among samples and identify possible contamination flows. ICU and NICU most abundant HAIrB-related oligotypes were visualized as frequency heatmaps (Figure 6). In ICU (Figure 6a), the most frequent oligotype detected was oligotype_1, classified by our pipeline as *S. epidermidis*, and present in ∼70% or more environmental samples, as well as in ∼50% of patient samples. Oligotype_4, identified as *S. aureus*, was also shared across environmental and patients samples in high proportions, as well as *P. aeruginosa* oligotype_12, *K. pneumoniae* oligotypes_19, _20, _6 and _63, *E. coli* oligotype_2, *E. faecium* oligotype_24 and *E. faecalis* oligotype_5.

**Figure 6.**
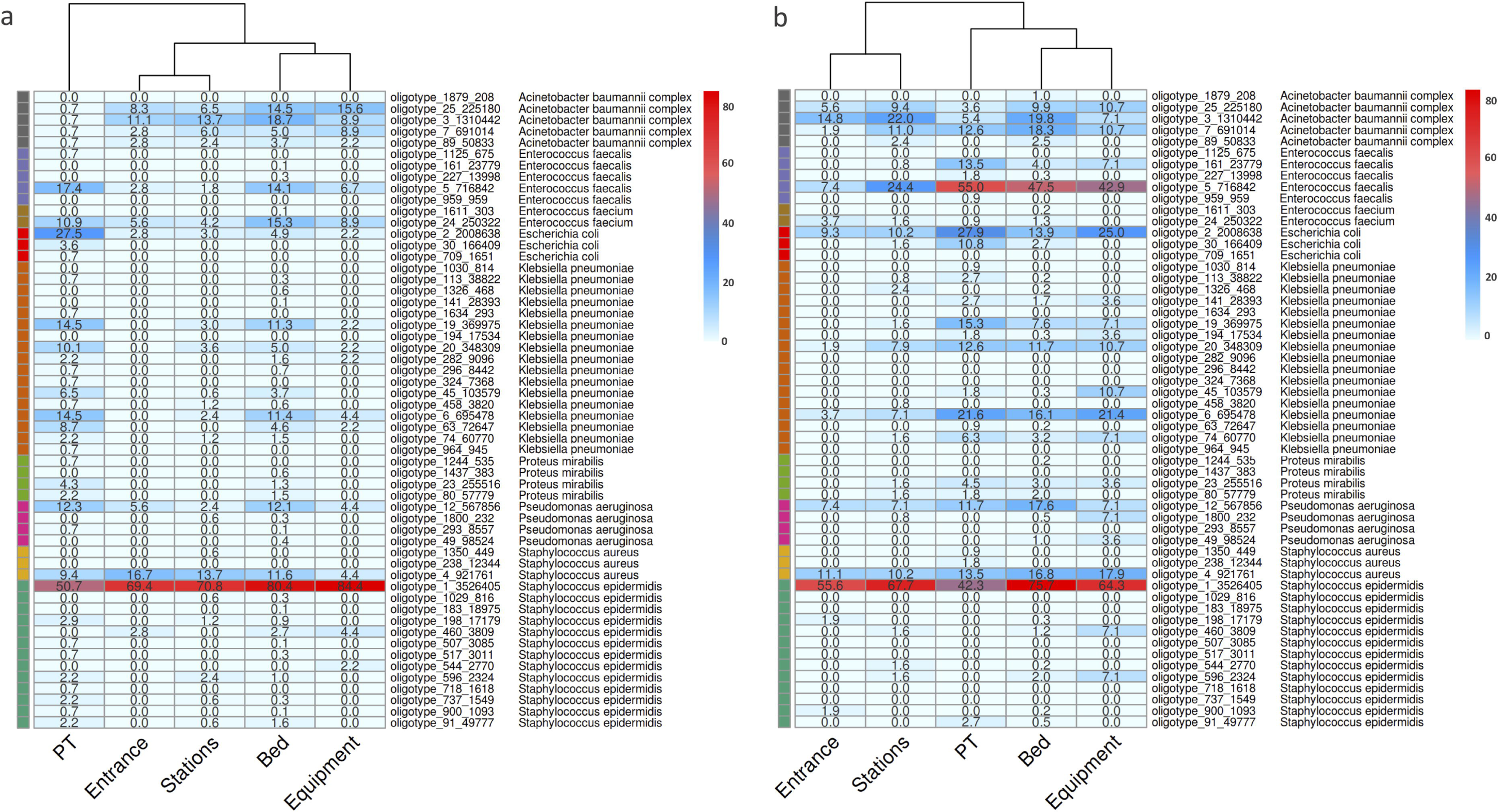
Source tracking. Oligotype heatmaps showing correspondence of their distribution across the sample locations. Top abundant HAIrB bacteria and their oligotypes were selected for visualization. (**A**) ICU and (**B**) NICU proportions of samples with the oligotype detected. Oligotypes are identified by a unique code, along with its count number (total reads detected along the analysis) and the bacterial taxonomy attributed by the Neotools pipeline. Color scales represents the proportion of positivity in the group of samples.

In general, the selected oligotypes showed highest proportions in samples from patients and beds, regardless of the unit sampled. Still, NICU samples seem to present higher dispersion. Except for samples from the hospital entrance, all the above cited oligotypes were found in elevated proportions in patient, bed, stations, and equipment samples. *A. baumannii* complex oligotypes (_7, _3, _25 and _89) were more abundantly found in healthcare environment samples. In NICU (Figure 6b), the most frequent oligotype in all sample locations was also *S. epidermidis* oligotype_1. *E. faecalis* oligotype_5 was abundantly found in patient, bed and equipment samples, showing lower but relevant presence in stations and entrances as well. Similar to ICU oligotypes, *E. coli* oligotype_2, *P. aeruginosa* oligotype_12, *S. aureus* oligotype 4 and *K. pneumoniae* oligotypes_19, _20 and _6 were highly dispersed and shared between different location samples. In NICU, *A. baumannii* complex oligotypes were detected in both patient and environmental samples, differing from ICU where they were prevalent in environmental samples.

The longitudinal profiles of the main oligotypes and their correspondent bacteria in ICU showed that oligotype_1 appears as the main source of *S. epidermidis* throughout the year, in the environment and in the patient samples (Supplementary figure 4a and 4b). A trend line from quantile regression suggests an abundance increase of this oligotype for patients from Februrary-2019, despite lower levels in June-2019. Other ICU HAIrB did not show specific oligotype-related temporal patterns, as they presented more heterogeneous and discontinuous monthly variations (Supplementary figure 4 c-f). NICU samples presented the same pattern for *S. epidermidis*, related to the same oligotype_1 sequence, highly abundant across all study period in the environment (Supplementary figure 4 g-h). Patient samples showed more significant increases from April-2019. In addition, NICU shows high abundance of *E. faecalis* oligotypes, mainly represented by oligotype_5 in patient stool samples, but also contaminating the surrounding environment - mainly the beds.

Detailed oligotype positivity rates across time points is shown in Supplementary figure 5. In addition, these contamination frequencies for HAIrB group were plotted as risk maps for ICU and NICU (Supplementary figure 6 and 7). These maps and red spots represent the monthly contamination in specific hospital locations, suggesting NICU has a more constantly contaminated environment than ICU, with no major HAIrB reduction over the year.

**Figure 7.**
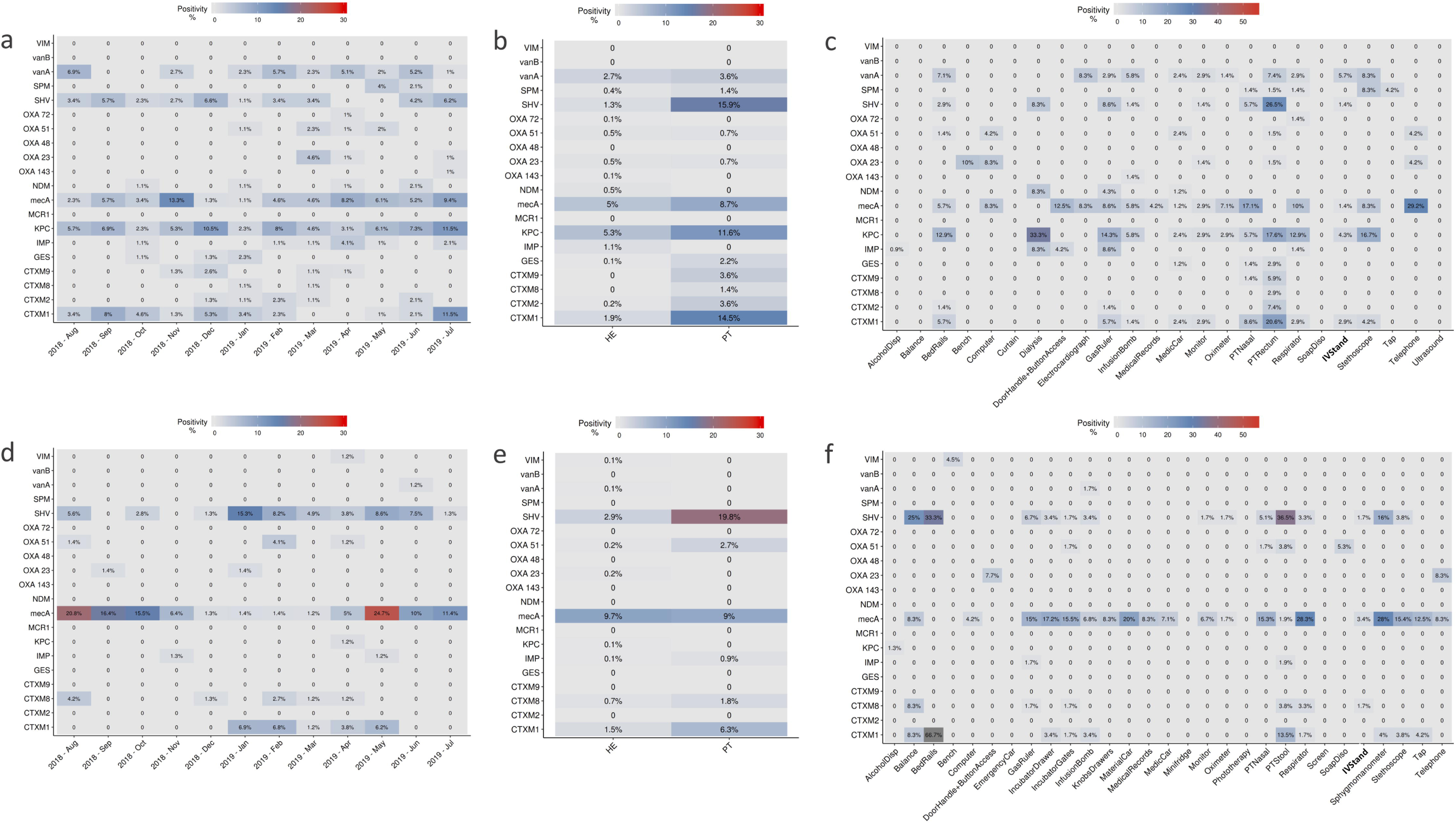
Antimicrobial resistance genes proportions of positivity in samples tested for (**A**) ICU along the year, (**B**) ICU between sources - patient (PT) and healthcare environment (HE), (**C**) ICU among specific sites, (**D**) NICU along the year, (**E**) NICU between sources - patient (PT) and environment (HE), (**F**) NICU among specific sites.

### Bacterial isolates identification and genomes characterization

ICU and NICU 17 bacterial isolates previously identified by the hospital microbiology laboratory using conventional techniques were analyzed through whole genome sequencing (WGS) and confirmed 88% of the previous microbiological results (15 out of 17 isolates) (Table 1). For the two diverging samples, one was identified just as *Klebsiella* by the microbiology lab but with WGS we were able to classify it as *K. pneumoniae*. Another sample, identified as *K. oxytoca* by the microbiology laboratory, was classified as *K. michiganensis* by WGS analysis. Additionally, 16S rRNA gene sequencing was performed for these isolated bacteria, also resulting in 88% agreement with the conventional microbiology laboratory, differing for the same two samples of *Klebsiella*. One of them (*Klebsiella*) was classified as *K. pneumoniae* by our bioinformatics pipeline and the other one (*K. oxytoca*) could not be differentiated below Enterobacteriaceae family level. Thus, WGS and 16S rRNA sequencing presented 94,11% agreement in the taxonomical classifications, only differing in the resolution level for the *K. michiganensis* in the amplicon 16S rRNA gene sequencing (Table 1).

Genomic investigation analysis was performed by searching for the specific 16S rRNA gene sequences in each isolated WGS results and comparing them to the oligotypes identified in the ICU and NICU amplicon samples. The same 16S rRNA fragment sequence corresponding to oligotype_4 was found in all *S. aureus* isolated genomes, even the one from before the project period. In the *S. epidermidis* WGS results we found the oligotype_1 sequence; *E. faecalis* presented the oligotype_5; *P. aeruginosa* oligotype_12; and some of the *E. coli* isolates had the oligotype_2 (Table 1). Two *E. coli* bacterial isolates from NICU in 2019 did not have any correspondence of their 16S rRNA sequences in the amplicon hospital microbiome survey. Furthermore, specific 16S rRNA sequences from *Klebsiella* WGS results, isolated from ICU and NICU, were not detected in the hospital microbiome oligotypes either.

In addition, genomic MLST profiling was performed along with clonality analysis using WGS results. These analyses separated *S. aureus* in three MLST strains (ST398, ST2383 and ST1435) and two clonal groups (*S. aureus* Group 1 and 2). Group 2 includes bacterial isolates from October-2018 and January-2019, both from NICU, however, these two isolates were more distantly related (195 SNVs, 99.93 ANIm and an average of 98.47% nucleotides aligned) than the Group1 isolates from October-2018 (97 SNVs, 99.99 ANIm and an average of 99.90% nucleotides aligned). The most different *S. aureus* (ST1435) was the only one carrying a *mecA* resistance gene and isolated in the hospital prior to this project. In addition the *S. epidermidis* isolated from the ICU in November-2018 belongs to ST2 and also carries the *mecA* resistance gene. The two *E. faecalis* were from the same MLST strain (ST21), but not sufficiently similar to be considered clonal (>5,000 SNVs, 99.59 ANIm and an average of 92.18% nucleotides aligned). One *E. faecalis* was isolated from ICU and the other from NICU. *P. aeruginosa* isolated, both from ICU, were identified as different MLST strains (ST245 and ST1816* containing 1 SNP from the canonical strain) and not clonally related (>9,000 SNVs, 99.29 ANIm and an average of 90.29% nucleotides aligned). Only two *E. coli* isolated from NICU in January-2019 belongs to the same MLST strain (ST69) and clonal group (*E. coli* Group 1 - with 10 SNVs, 99.97 ANIm and an average of 99.74% nucleotides aligned). However, these clonal *E. coli* genomes are the ones without corresponding 16S rRNA oligotypes in the microbiome survey. This WGS analysis from bacterial isolates corroborates our bioinformatics pipeline taxonomical identification for the species-level classification of most relevant oligotypes identified in this microbiome project.

### Antimicrobial resistance genes profile in the hospital microbiome samples

Antimicrobial resistance genes were searched every month in healthcare environment and patient samples in both ICU and NICU. In ICU, a total of 135 environmental samples (14%) and 51 patient samples (36%) were positive for any resistance gene. In 3.7% of environmental samples and 15% of patient samples more than one resistance gene was detected. In 72 ICU beds tested, 22 of them showed the same resistance gene found in samples from its respective inpatient. Resistance genes positivity over the twelve-month period in the ICU showed that *mecA, bla*_CTX-M-1 group_, *bla*_SHV-like_, *bla*_KPC-like_ and *vanA* were the most prevalent genes detected (Figure 7a). Other genes were detected in lower frequencies and in different periods, such as *bla*_SPM-like_ detected from May to June – 2019, *bla*_OXA-51-like_ detected from January to May – 2019, and *bla*_NDM-like_ not continuously detected. Patients showed more positive samples than healthcare environment (Figure 7b). However, proportionally, patients have far fewer samples collected than environment. In terms of specific sampling sites (Figure 7c), *mecA* gene was found widely distributed and with higher proportions in samples such as patient nose and equipment as telephone, door handles/access buttons, respirators, computers, electrocardiograph, gas ruler, oximeters, among others. *bla*_KPC-like_ was frequently found in dialysis equipment, stethoscope, gas ruler and bedrails, as well as in patient rectum samples and similar to *bla*_SHV-like_ distributions. *vanA* gene distributions is also higher in stethoscopes, electrocardiograph, bedrails, infusion bomb and patient rectum samples.

In NICU, a total of 112 environmental samples (13%) and 32 patient samples (30%) tested positive for any resistance gene. In 1.6% of environmental samples and 9.9% patient samples more than one resistance gene was detected. Among 60 NICU tested beds, 19 showed the same resistance genes as its respective inpatient. Assessment of AMR genes in the NICU demonstrated higher positivity of *mecA* gene over all the twelve months, with higher frequencies from August-2018 to November-2018 and from May-2019 to July-2019 (Figure 7d). *bla*_CTX-M-1 group_, *bla*_SHV-like_ were also detected, but most frequently from January-2019 to June-2019. Patients also showed high positivity for AMR genes, with exception for *mecA* gene, which was even more present in environmental samples (Figure 7e). Also, *mecA* gene was found in higher proportions of most samples analyzed (Figure 7f), but even more frequent in respirators, sphygmomanometers, material cars, incubators gates and drawers, gas ruler and stethoscopes samples. *bla*_SHV-like_ was mostly found in patient stool samples as well as in balance, bedrail and sphygmomanometer samples.

Correlating the most abundant AMR genes in patients with healthcare environments, and adjusting the proportion of samples for each group, we could observe that there is an overlapping frequency pattern: when the AMR gene is in higher frequencies for environmental samples, they are also higher in patient samples (Figure 8). Giving the experimental design carried out in this study, we were not able to define the actual contamination source, the environment or the patient. We could, nonetheless, observe their close relationship.

**Figure 8.**
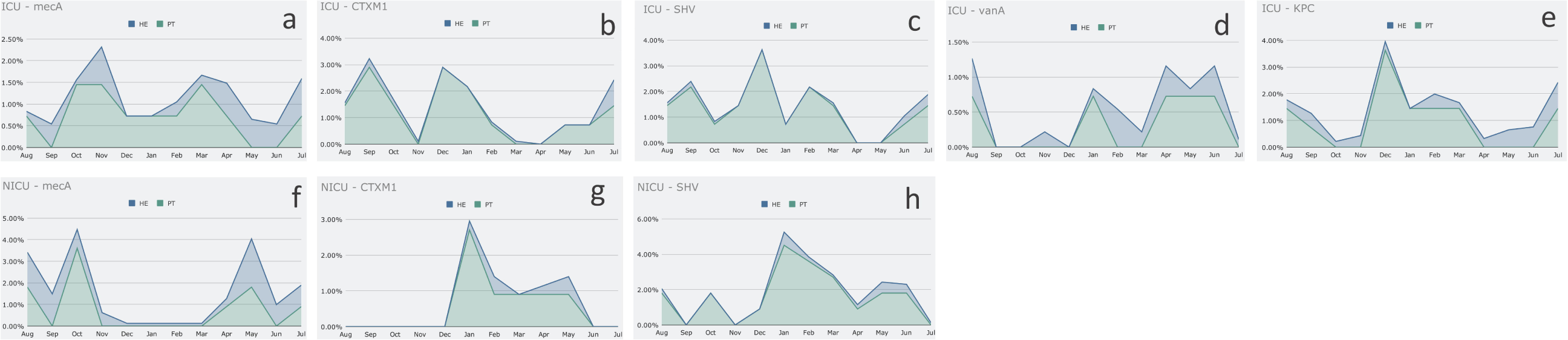
Antimicrobial resistance gene frequencies in patients (PT) and healthcare environment (HE) for each month analyzed across the year. (**A**) *mecA* gene in ICU, (**B**) *bla*_CTX-M-1_ group gene in ICU, (**C**) *bla*_SHV -like_ gene in ICU, (**D**) *vanA* gene in ICU, (**E**) *bla*_KPC -like_ gene in ICU, (**F**) *mecA* gene in NICU, (**G**) *bla*_CTX-M-1_ group gene in NICU, (**H**) *bla*_SHV -like_ gene in NICU.

## Discussion

Hospital microbiome comprehension, and more specifically bacterial pathogen tracking, can be of great relevance to understand, reduce and prevent healthcare associated infections (HAI). In this paper we have demonstrated the application of high-throughput, next generation (NGS) amplicon sequencing to screen, monitor and track bacterial profiles in a hospital environment. Additionally, we used the same NGS samples for AMR/MDR genes profiling through real-time PCR and compared the amplicon results with WGS analysis of bacterial isolates from ICU and NICU.

Several bacterial investigations have been carried out in intensive care units, since ICUs are related to high rates of HAI and MDR acquired infections (4,21,41–43). Some ICU studies using high-throughput sequencing reveal bacterial profiles that would not be recovered only by conventional microbiology methods, given its specificity and selectivity for known types of bacteria (5,44,45). The method we used in this study was also culture-independent, but designed to allow comparisons relative to the bacterial load in the samples. Previous studies have already demonstrated that total sequence reads from NGS samples (library size) do not need to be arbitrary, thereby allowing bacterial load estimation (24,46). Thus, allowing the identification of temporal profiles and critical sample collection sites suited to be the focus of hospital cleaning interventions.

The bacterial profile found in this study for ICU and NICU environments is similar to other intensive care studies, with Proteobacteria and Firmicutes as the most abundant bacterial phyla detected (1,2,5). However, most abundant bacterial genera and species detected in our study varied, particularly because of the high rates of specific HAI related bacteria detected.

The overall results show that the hospital environment maintains a close relationship with the patients’ microbiome in their respective care units, a pattern also observed for AMR genes. In this project, the experimental design was not constructed to identify the definitive sources of bacterial contaminations (patients, environment or healthcare professionals), as done by other studies that could correlate the contamination of patients according to the environment and their length of stay at the hospital (1). However, we observed the NICU bacterial profiles with wide and continuous contaminations over the year, suggesting a flow between patients and environment by factors like healthcare workers or visitors, since NICU patients are restricted to their incubators and do not have deliberated access to their surrounding environment. Other studies already showed the resemblance between NICU environment and the gut microbes of premature infants (47). Some sample positivity reduction observed in NICU for *S. aureus, S. epidermidis* and *mecA* AMR gene could be attributed to infection control interventions during November and December of 2018. The NICU underwent exhaustive cleaning and sanitation processes, including shifting locations from November of 2018 until January 2019. In the ICU rigorous hygienization processes were also reported by the hospital in December-2018. This could explain the variations in the bacterial and AMR profiles during this period and shortly after.

In both ICU and NICU, throughout this study period, patient samples yielded the highest bacterial load, despite their lower alpha-diversities. Yet, these comprised the only sample group with apparent clustering in beta-diversity analysis. This patient profile can be explained by the fact that they showed a particular microbiota, with predominance of only a few bacteria, which is generally expected for nasal samples, but not for fecal/rectal samples (48,49). This diversity may be related to patient infections or microbiota imbalance due to hospitalization (50–52). Recent studies have shown that immediately after birth, newborns are colonized by the maternal and surrounding microbiota. Infants from cesarean sections, mothers treated with prophylactic antibiotics, or not breastfed during the neonatal period have altered microbiota profiles, as well as colonization by opportunistic pathogens associated with the hospital environment, such as *Enterococcus, Enterobacter* and *Klebsiella* (Shao et al., 2019). The implications of microbiota changes during hospitalization period is not fully understood yet. Another healthcare issue that may be directly associated with microbiome diversity is the development of sepsis. Studies have identified low intestinal microbiota diversity and *Staphylococcus* predominance as risk factors for sepsis in neonates born at 24-27 weeks of gestation (53). In adults, the risk of sepsis has also been studied related to the microbiome profile imbalance and other associated factors (54).

We detected AMR genes more frequently in rectal/stool samples, but also considerable in nasal samples. *mecA, bla*_CTX-M-1 group_, *bla*_SHV-like_ and *bla*_KPC-like_ were by far the most frequent AMR genes detected in patients, but also spread in the hospital environment. Meticillin-resistant *S. aureus* (MRSA) carrying *mecA* gene is a rising threat to public health (55). This gene has been found not only in hospital environments, but also circulating in the community and not only restricted to *S. aureus*, but also detected among other species from the genera, including *S. epidermidis* (56,57). Carbapenem resistance and Extended spectrum β-lactamases genes detected here (*bla*_KPC-like_, *bla*_CTX-M-1 group_, *bla*_SHV-like_ and) were commonly reported in studies from Brazilian hospital environments as a serious problem in urgent need of actions to reduce their spreading rates (58,59).

In both NICU and ICU units, we highlight the large variation across monthly assessed specific sites. In each month, while the overall microbial load varied in one direction (increase or decrease), there were still samples indicating divergent degrees of contamination. This suggests a lack homogeneity or reproducibility in the sanitation processes. Another possibility, the rapid re-contamination of the environment with HAI bacteria after cleaning due to external sources, e.g., healthcare workers and visitors. This study was not designed to evaluate the sanitation processes, but rather to deepen our understanding of the hospital bacteriome. WGS analysis of bacterial isolates allowed us to complement the amplicon sequencing results by confirming the taxonomical identification for several 16S rRNA amplicons and by showing that bacterial microbiome profiles, recovered in patient and environmental samples, were from real significant bacterial strains widely spread in the hospital intensive care units.

Therefore, through a one-year-long hospital surveillance study we were able to demonstrate how NGS technologies can be applied to large-scale, fast, and cost-effective monitoring of hospital microbial contamination, thereby improving infection control practices. Effective identification of HAI hotspots and contamination flows with pathogenic bacteria, in multiple sampling sites simultaneously, represents an unprecedented resolution gain for hospital microbiological control and epidemiological surveillance.

## Data Availability

All sequence data are deposited in NCBI BioProject PRJNA604445

## Author Contributions

AC performed the analysis and wrote the manuscript. AS, GC, LO performed the analysis and reviewed the manuscript. DB, VS performed the laboratory sample analysis. AN, AR, RV, AM, BB, TR performed the sample collection. AS, LO, CH, TR, MM, FM were responsible for the project design and approvals.

## Data availability

All sequence data are deposited in NCBI BioProject PRJNA604445

## Supplementary figures

**Supplementary Figure 1**. Shannon diversity profiles.

**Supplementary Figure 2**. Weighted UniFrac Beta-diversity profiles for all samples collected in both units, ICU and NICU separated in three different timelines.

**Supplementary Figure 3**. Most abundant bacteria detected and classified in four taxonomical ranks.

**Supplementary Figure 4**. Longitudinal profile of specific bacteria and its oligotypes most abundantly found in ICU and NICU.

**Supplementary Figure 5**. Proportion of positive samples for the most abundant oligotypes in ICU and NICU.

**Supplementary Figure 6**. Risk map ICU.

**Supplementary Figure 7**. Risk map NICU.

**Supplementary table 1**. Sample collection sites

**Supplementary table 2**. Monthly collected samples and sequencing information.

